# Real-world safety of zonisamide: Mining and analysis of adverse events related to zonisamide based on FAERS database

**DOI:** 10.1101/2024.12.04.24318510

**Authors:** Yongyi Zhang, Zhongqian Sun, Haoming Li, Qingxia Kong, Xuezheng Zhang, Chengde Li

**Author notes:** Correspondence author : E-mail addresses (Chengde Li) (Xuezheng Zhang). These authors contributed equally to this work and should be considered co-corresponding authors (Xuezheng Zhang, Chengde Li). **Ethics approval** Anonymized data were collected from a publicly available database and do not require approval from the ethics committee. **Availability of data and material** Data are available on the FAERS database. Analytical datasets are available from the corresponding author upon reasonable request. **CRediT Authopship Contribution Statement** Chengde Li: Conceptualization, Methodology, Validation, Writing- Reviewing and Editing Yongyi Zhang: Formal anlgysis, Data curation, Writing- Original draft preparation, investigation. Zhongqian Sun: Data curation, Formal anlgysis, Writing- Original draft preparation. Haoming Li: Data curation, investigation. Qingxia Kong: Resources, Funding investigation. Xuezheng Zhang: Conceptualization: Methodology, Validation,Writing- Reviewing and Editing All authors have read and approved the final manuscript. **Declaration of Competing Interest** The authors declare that they have no known competing financial interests or personal relationships that could have appeared to influence the work reported in this paper.

## Abstract

**Purpose:** To investigate the real-world adverse event signals associated with zonisamide and provide a foundation for its safe clinical use.

**Methods:** Adverse event reports involving zonisamide as the primary suspected drug were collected from the FDA Adverse Event Reporting System (FAERS) database, covering the period from the first quarter of 2004 to the fourth quarter of 2023. The data were analyzed utilizing the Reporting Odds Ratio (ROR) and Bayesian Confidence Propagation Neural Network (BCPNN) methods of the proportional imbalance technique.

**Results:** A total of 3205 adverse event reports involving zonisamide as the primary suspected drug were identified, resulting in 260 positive signals for preferred terms (PTs). These signals, derived from both the ROR and BCPNN methods, encompassed 27 systems and organs (SOCs), with a predominant focus on nervous system disorders and skin and subcutaneous tissue disorders. The most frequently reported PTs included seizures, drug reactions with eosinophilia and systemic symptoms, Stevens-Johnson syndrome, rash, and dizziness. Notably, the top PTs in terms of signal intensity included perinephritis, epilepsy with myoclonic-atonic, ocular mucocutaneous syndrome, ocular choroidal leakage, tonsillar exudate, and ovarian granulosa vesicular cell tumor. Interestingly, ten of the top 30 risk signals for adverse events, based on signal strength, were not detailed in the package inserts. Thses included perinephritis, myoclonic dystonic epilepsy, ovarian granulosa vesicular cell tumor, positive human herpesvirus 6 serology, and positive lymphocyte stimulation test.

**Conclusion:** Common adverse reactions to zonisamide in real-world settings are generally in line with the established specification, with the most frequently observer signals related to neurological, skin, and subcutaneous tissue disorders. However, several newly suspected adverse reactions have been identified, including perinephritis, infectious pneumonitis, ovarian granulosa vesicular cell tumor, positive serology for human herpesvirus 6, and positive lymphocyte stimulation test. These findings indicate that these potential adverse reactions should be closely monitored in clinical practice.

## 1. Introduction

Zonisamide (ZNS), chemically known as 1,2-benzoxazole-3-methanesulfonamide, is a novel antiepileptic drug characterized by its diverse mechanisms of antiepileptic action. It blocks voltage-dependent Na+ channels and T-type Ca2+ channels, while also potentiating γ-aminobutyric acid-mediated inhibition and reducing glutamate-induced excitability [1,2]. It was approved by the FDA in 2000 for the treatment of adult focal epilepsy, and was approved as add-on therapy for adult focal seizures in China in 2009. In recent years, in addition to its antiepileptic effects, zonisamide has been gradually found to have multiple pharmacological effects, such as inhibition of oxidative stress, neuroprotection, and inhibition of microglia activation.These attributes have led to improved therapeutic outcomes in various clinical applications, such as Parkinson’s disease, idiopathic tremor, migraine, neurological headache, obesity, and mood disorders,. [3,4]. Notably, in January 2009, zonisamide was also approved in Japan for the treatment of Parkinson’s disease.

With the increasing indications and clinical applications of zonisamide, adverse drug reactions have become a significant concern. A multicenter, prospective, randomized controlled, open-label, non-inferiority trial demonstrated that the incidence of adverse drug reactions to zonisamide was as high as 45%, with serious adverse reactions occurring in 1.22% of cases [5]. Additionally, a cohort study examining epilepsy cases in China indicated that the most common adverse reactions to zonisamide included loss of appetite (11.8%), dizziness (6.9%), and headache (3.9%) [2]. Adequate recognition of zonisamide’s adverse drug reactions, early identification of high-risk factors, and effective management strategies can prevent or mitigate the occurrence of these reactions, thereby enhancing the overall management of patients with epilepsy.

Despite many years of clinical use of this drug for epilepsy, there remains a need to gather additional information for safety assessment, particularly given the limited experience with its clinical use in new indications such as Parkinson’s disease, migraine, and obesity. Furthermore, there is a notable lack of systematic studies examining the adverse drug events (AEs) associated with ZNS based on real-world data. The FDA Adverse Event Reporting System (FAERS) serves as a crucial pharmacovigilance database that collects spontaneously reported AEs from healthcare professionals, pharmaceutical manufacturers, patients, and legal representatives across various regions worldwide. It has been extensively utilized for screening drug safety information. In this study, we conducted an AE risk signal mining analysis of ZNS using data from the FAERS database to investigate the occurrence of AEs associated with ZNS in real-world settings, aiming to provide valuable insights for the safe clinical use of ZNS.

## 2. Materials and methods

### 2.1 Data source and processing

Adverse event reports from the FARES database (https://fis.fda.gov/extensions/FPD-QDE-FAERS/FPD-QDE-FAERS.html) were collected from Q1 2004 to Q4 2023 and searched using specific terms related to the drug Zonisamide (ZONISAMIDE, ZONEGRAN, EXCEGRAN, TRERIEF). Duplicate reports were removed based on unique patient identifiers (primaryID), incomplete reports were excluded.The data were processed to focus specifically on adverse event reports in which Zonisamide was the primary suspected drug.

### 2.2 Methods

#### 2.2.1 Data processing

The Medical Dictionary for Regulatory Activity (MedDRA 26.1) was used in this study to standardize all PTs, which were then mapped to system organ classification (SOC) for further analysis. Doubtful and blank data, were also removed to form the raw data for this study.

#### 2.2.2 Statistical analysis

Data on patients’ gender, age, prognosis, and AE occurrence were extracted from AE reports for descriptive statistical analysis. Reporting Odds Ratio (ROR) and Bayesian confidence propagation neural network (BCPNN) were used to jointly detect the AE risk signals of ZNS, and both methods were based on the proportional imbalance measure (refer to Table 1),detection of potential AE risk signals by comparing the proportion of target events occurring for the target drug with the proportion occurring at the current time for all other drugs in the same period,where A is the number of targets AE of the target group, B is the number of other AEs of the target group, C is the number of target AE of other groups, and D is the number of other AEs of other groups. The calculation formulae and positive signal criteria for the 2 methods are shown in Table 2. The signal values of all selected PTs of zonisamide were calculated to analyse the occurrence of their associated AEs, and then the PTs were classified according to the SOC in MedDRA version 26.1. The generation of AE signals suggests the existence of a statistical association with the drug, and the higher values of the signals represent the stronger associations. The SAS 9.4 software was used for data processing and analysis.

**Table 1.** Four-fold table of disproportionality measurement.

|  | Number of<br>target<br>adverse<br>reaction reports | Number of other<br>adverse<br>reaction reports | Total |
| --- | --- | --- | --- |
| Target drugs | A | B | A+B |
| Other drugs | C | D | C+D |
| Total | A+C | B+D | N=A+B+C+D |

**Table 2.**
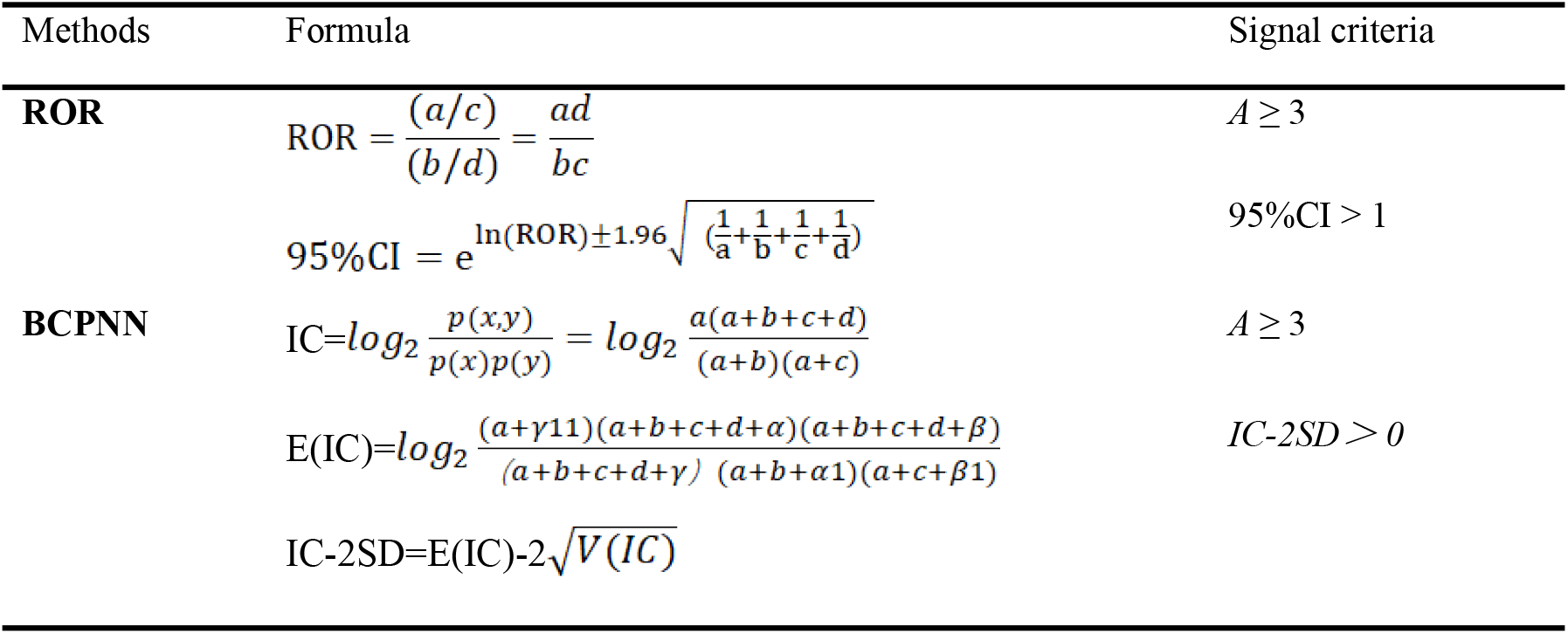
Measure of Disproportionality and signal generation criteria.

In this study, those with suspicious AE risk signals in both methods were identified as zonisamide AE risk signals, and the new risk signals were screened against the ZNS FDA specification, the European Medicines Agency specification and the Japanese specification.

## 3 Results

### 3.1 Zonisamide-related AE Reporting Basics

A total of 17,307,196 AE reports were received in the FAERS database from Q1 2004 to Q4 2023, and 3,205 AE reports were received with ZNS as Primary suspect drug. The basic information of the patients involved, such as gender, age, country/region, reporter, primary disease, and regression, is shown in Table 3.

**Table 3.** Characteristics of AE reports associated with Zonisamide.

| Charcateristics | N | Propotion(%) |
| --- | --- | --- |
| **Total** | 3205 | 100.00 |
| **Gender** |  |  |
| Female | 1335 | 41.65 |
| Male | 1020 | 31.83 |
| Not Specified | 850 | 26.52 |

| Charcateristics |  | N | Propotion(%) |
| --- | --- | --- | --- |
| Age (year) | <18 | 372 | 11.61 |
|  | ≥18, <45 | 723 | 22.56 |
|  | ≥45, <65) | 427 | 13.32 |
|  | 65≤ | 357 | 11.14 |
|  | NotSpecified | 1326 | 41.37 |
| Report countrier | United States of America | 1245 | 38.85 |
|  | Japan | 921 | 28.74 |
|  | United Kiongdom | 228 | 7.11 |
|  | France | 127 | 3.96 |
|  | Germany | 126 | 3.93 |
| Degree of seriousness | Serious | 2773 | 86.52 |
|  | Non-Serious | 432 | 13.48 |
| Outcome | Life-Threatening | 189 | 5.90 |
|  | Hospitalization | 1220 | 38.07 |
|  | Disability | 111 | 3.46 |
|  | Death | 192 | 5.99 |
|  | Other | 1510 | 47.11 |
| Time of occurrence | 0-30d(%) | 601 | 18.75 |
|  | 31-60d(%) | 174 | 5.43 |

### 3.2 SOC profile of zonisamide-associated AE signalling involvement

The analysis of 3205 AE reports with zonisamide as the primary suspected drug included a total of 9804 adverse events, with 504 preferred terms (PTs) ultimately being analyzed. Among these, 299 positive PT signals were identified using the ROR method and 260 positive PT signals were identified using the BCPNN method. A total of 260 positive PT signals, met the criteria of both methods. Additionally, these signals involved 27 system organ classes (SOCs). The distribution of the number of PT signals and AEs under each SOC is illustrated in Figure 1. The top 5 System Organ Classes with the highest number of Preferred Terms were nervous system disorders, psychiatric disorders, investigations, General disorders and administration site conditions, as well as skin and subcutaneous tissue disorders. On the other hand,, the top 5 SOCs with the highest number of Adverse Events (AEs) reports were nervous system disorders, General disorders and administration site conditions, psychiatric disorders, skin and subcutaneous tissue disorders, as well as injuries, poisonings, and procedural complications.

**Figure 1.**
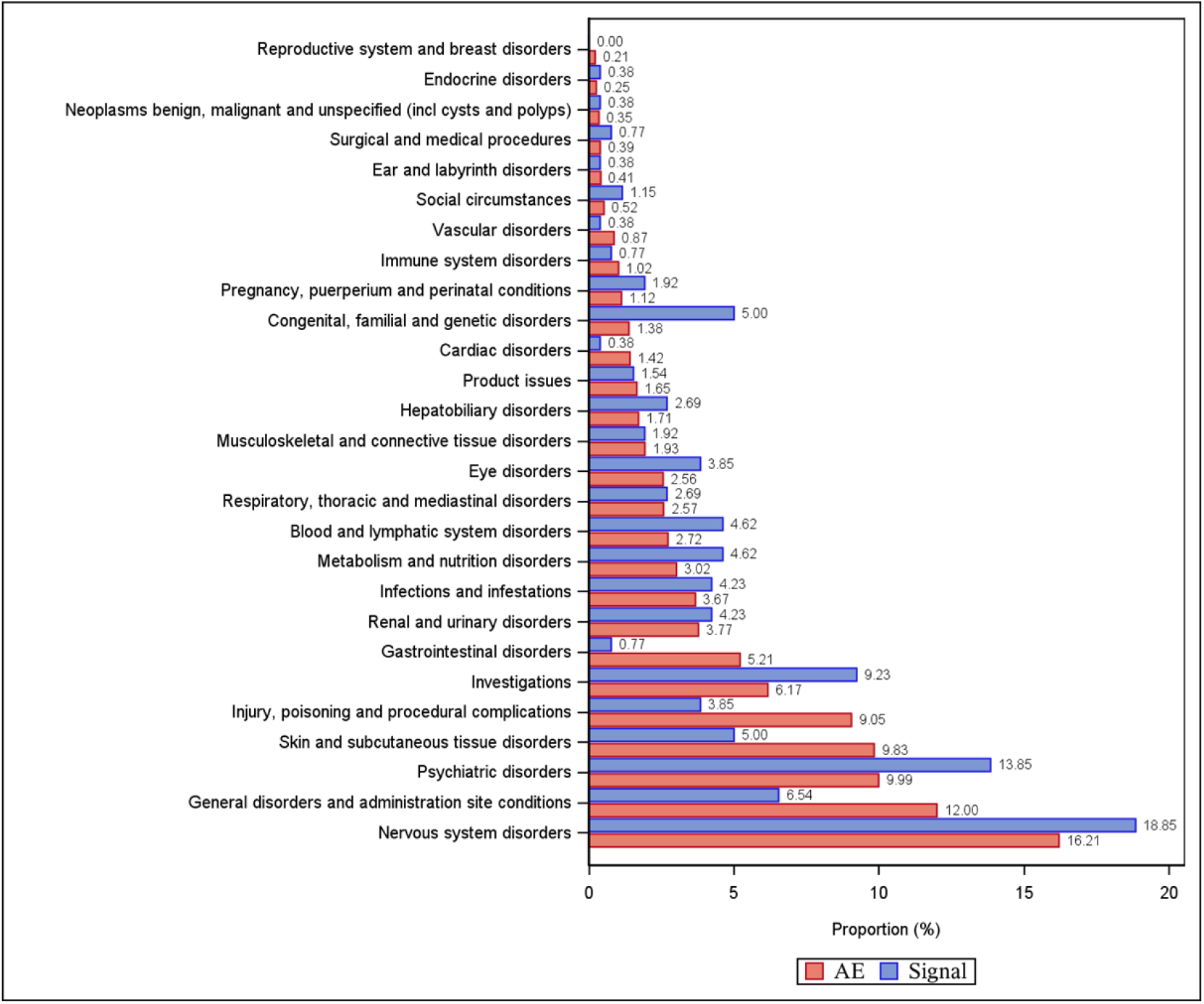
Positive signals and Adverse events of zonisamide-involved system organ lassification in the FEARS database.

### 3.3 Risk Signal Analysis for AE associated with Zonisamide

The 260 risk signals that were positive in both ROR and BCPNN methods were ranked in descending order based on frequency of occurrence. Table 4 displays the top 30 ADR signals in terms of frequency. Among the top 6 risk signals by frequency were epileptic seizure, drug reaction with eosinophilia and systemic symptoms, Drug ineffective, Stevens-Johnson syndrome, rash, and dizziness, primarily associated with Nervous system disorders, Skin and subcutaneous tissue disorders. Notably, two of the top 30 ADR risk signals were not documented in the package inserts, namely infectious pneumonia and over-the-counter use.

**Table 4.**
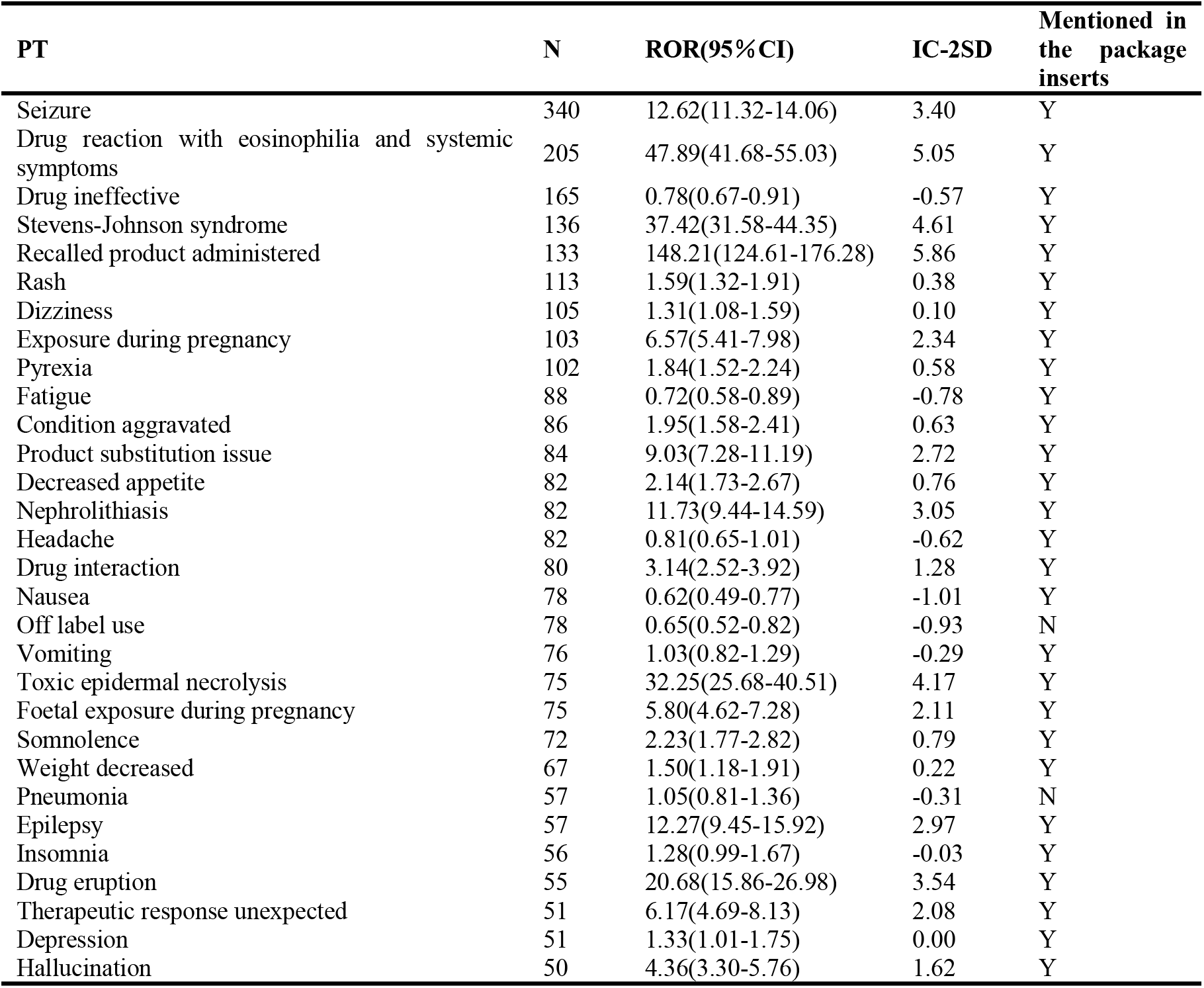
Top 30 risk signals for the frequency of zonisamide.

The 260 risk signals that were were identified as positive by both the ROR and BCPNN methods were ranked in descending order of signal intensity, and the top 30 AE signals are shown in Table 5. The six primary terms with the highest signal intensity included Perinephritis, Epilepsy with myoclonic-atonic seizures, Oculomucocutaneous syndrome, Choroidal effusion, Tonsillar exudate, and Ovarian granulosa-theca cell tumor. Notably, ten of the top 30 risk signals in terms of adverse drug reaction signal intensity were not previously documented in the package inserts, such as Perinephritis, Epilepsy with myoclonic-atonic seizures, Ovarian granulosa-theca cell tumor, Human herpesvirus 6 infection, Lymphocyte stimulation test positive.

**Table 5.**
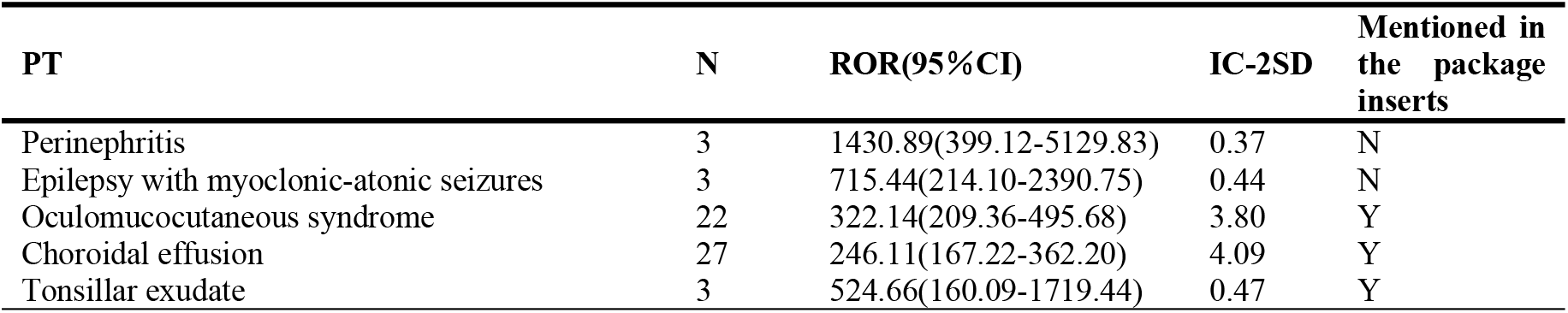
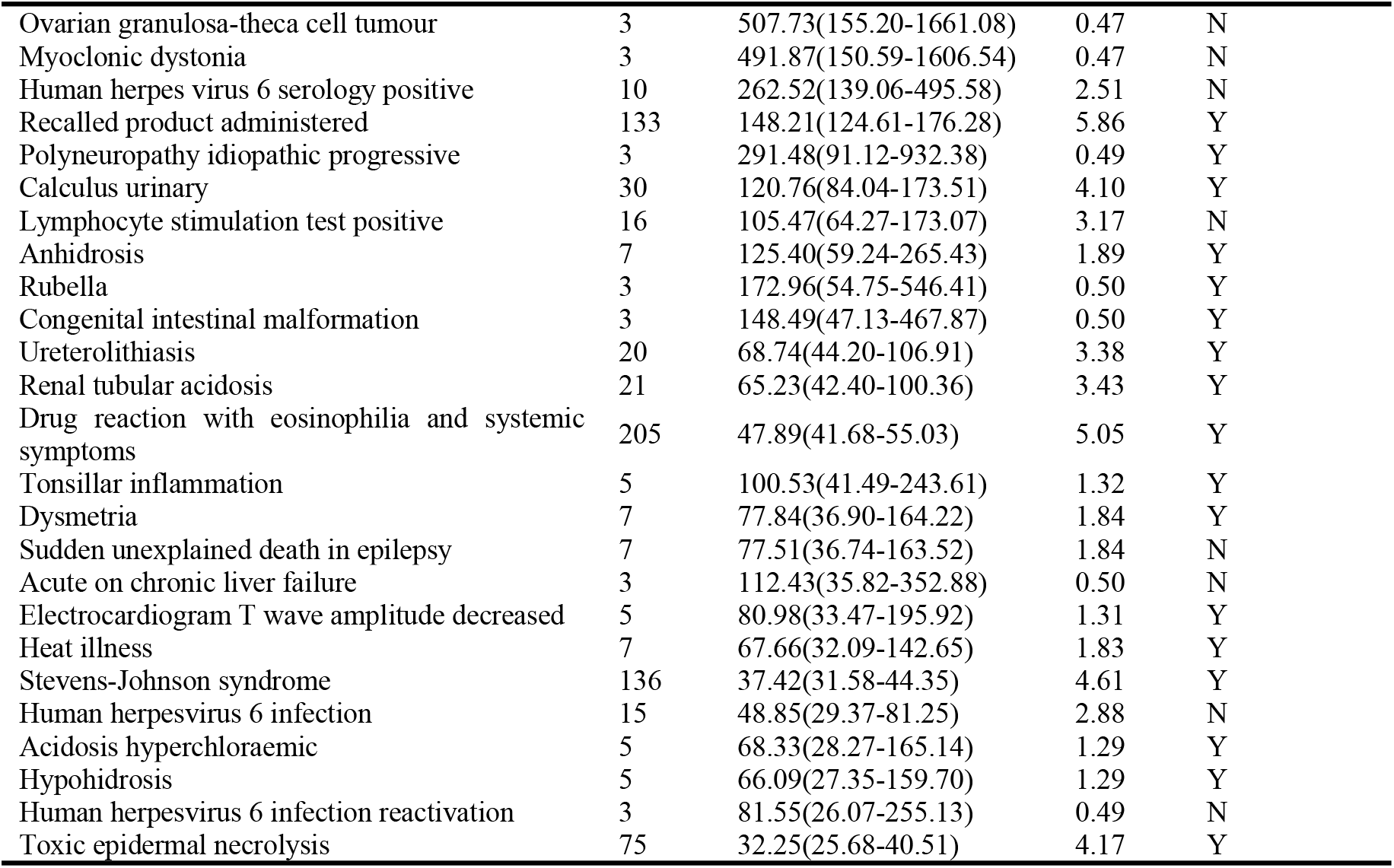
Top 30 risk signals for the signal intensity of zonisamide (sorted by ROR value)

## 4 Discussion

### 4.1 Basic information of Zonisamide AE Reporting

The study analyzed risk signaling of ZNS adverse events in the FAERS database from Q1 2004 to Q4 2023, revealing a total of 3,205 patients with zonisamide as the primary suspected drug. Females accounted for 41.65% of the population, slightly higher than males at 31.83%, contrary to the expected male-to-female gender ratio in epileptic morbidity (1.15-1.7):1[7]. Gender-specific studies on the occurrence of ZNS adverse events are lacking. ADRs were found to occur across all age groups with no significant differences in incidence, emphasizing the need for careful monitoring of zonisamide-treated cases to mitigate ADR risks.

The study found that the percentage of serious cases (86.52%) was significantly higher than non-serious cases (13.48%), with a higher percentage of fatal (5.99%) and disabled (3.46%) cases. Zonisamide, classified as a sulphonamide, can lead to serious and potentially fatal adverse reactions like Stevens-Johnson syndrome, toxic epidermal necrolysis, fulminant hepatic necrosis, granulocyte deficiency syndrome, and aplastic anaemia. Borrelli et al. examined adverse event reports in the antiepileptic drug FAERS and discovered that zonisamide has a high risk ratio for causing Stevens-Johnson syndrome / toxic epidermal necrolysis (ROR 70.2, 95% CI 33.1-148.7). This risk is 24.3 times higher than that of non-antiepileptic drugs[8], indicating that if a rash appears in patients treated with zonisamide, discontinuation of the drug at an early stage may help mitigate the frequency and severity of these adverse events.

### 4.2 SOC analysis of ZNS-associated AE risk signals

By analyzing real-world data, this study found that adverse events associated with ZNS were primarily linked to nervous system disorders (such as headache, dizziness, ataxia, somnolence, and seizures), psychiatric disorders (including insomnia, depression, hallucinations, and impulsivity), and skin/subcutaneous tissue disorders (such as rash, drug reaction with eosinophilia and systemic symptoms, Stevens-Johnson syndrome, and drug eruption). The adverse event identified in this study were generally in line with the descriptions provided in the drug documentation. It is crucial to raise awareness about these risks among healthcare providers and patients, particularly focusing on recognizing psychiatric and behavioral symptoms to mitigate the occurrence of aggressive behavior and suicide[9].

This study is the first to analyze safety information of ZNS using the FAERS database. The results indicate that the most prominent risk signals for adverse events are primarily related to skin and subcutaneous tissue disorders, such as Drug reaction with eosinophilia and systemic symptoms, Stevens-Johnson syndrome, Toxic epidermal necrolysis, and Oculomucocutaneous syndrome. Additionally, there are notable signals of neurological disorders, including dizziness, drowsiness, ataxia, epileptic futility, and seizures. Many of these signals align with information provided in the drug documentation highlighting the accuracy of signal mining in this study.

### 4.3 ZNS-related AE risk signal analysis

#### 4.3.1 Nervous system disorders - Seizure

In the SOC of nervous system disorders, specific adverse reactions, such as Myoclonic dystonias(ROR=491.87,IC_025_=0.47), epilepsy with myoclonic atonic seizures (ROR=715.44, IC_025_=0.44), seizure, showed high frequency and significant ROR values. Interestingly, these adverse reactions were not explicitly mentioned in the drug insert. This omission may be attributed to the fact that ZNS is often used as an add-on therapeutic agent in refractory epilepsy, where efficacy may not be significant or treatment may be unsuccessful [10,11], and several controlled studies involving the add-on therapy of zonisamide for focal epilepsy have shown that the seizure-free rate after the application of zonisamide ranges from only 6.2% to 18.1%[1,11]. Epilepsy with myoclonic atonic seizures ranked second in signal intensity. Myoclonic-atonic seizures are a refractory epilepsy syndrome, and it has been demonstrated that some sodium blockers such as carbamazepine, phenytoin sodium, and oxcarbazepine cause exacerbation of this epilpsy and even induce persistent status[12]. Whether zonisamide, a sodium blocker, also exacerbates this epilepsy requires further clinical studies[13]. In the context of the current study, zonisamide needs to be used with caution in patients with myoclonic-atonic epilepsy.

#### 4.3.2 Psychiatric disorders

The study revealed that zonisamide-induced psychiatric adverse events (AEs) accounted for 13.85% of reports, ranking as the second highest among System Organ Classes. Psychiatric symptoms mainly included delusions, irritability, hallucinations, suicide attempts, inattention, and memory difficulties. These cases were observed across all age groups, consistent with existing literature[10]. Most cases occurred within 1 month of treatment initiation. Zonisamide led to a higher incidence of irritability in adults (12.7%) compared to children (9.0%), as well as a higher incidence of depression in adults (13.9%) compared to children (2.3%)[15]. Adverse drug reactions in the psychiatric system typically manifest during the initial dose-escalation phase, with symptoms like irritability and depression resolving either spontaneously or upon discontinuation of the drug. However, speech abnormalities may persist, even after one year of treatment. In a prospective randomized study, 47% of patients reported dose-dependent speech abnormalities at one year, primarily involving impaired verbal long-term memory, verbal fluency, and mental dexterity, particularly at doses of 300 mg per day or higher[16]. It is recommended to systematically monitor cognitive function to enhance treatment outcomes.

#### 4.3.3 Renal and urinary disorders-Calculus urinary

The study revealed that the signal intensity ROR values and IC-2SD values were higher for renal and urological diseases such as renal stones, ureteral stones, and renal tubular acidosis, Perinephritis was identified as particularly noteworthy. Long-term observations showed symptomatic renal stones in 1.2-1.4% of patients and asymptomatic renal stones in 2.6% [14]. The incidence of renal stones was 2.87% within the first 6 months of treatment, 6.26% between 6 to 12 months, and 2.43% after 12 months. Zonisamide can lead to severe urinary adverse drug reactions, especially in individuals with renal insufficiency or those taking high doses of the medication [17]. Zonisamide exhibits a mild carbonic anhydrase inhibitory effect[18], potentially contributing to electrolyte imbalance-related adverse reactions. Moreover, ZNS may elevate urinary sodium, calcium (hypercalciuria), and oxalate excretion, as well as urinary concentration, promoting the formation of calcium phosphate stones. This, in turn, can exacerbate kidney damage and dysfunction in severe cases [19].Increasing fluid intake and promoting urine excretion can mitigate the risk of kidney stone formation, particularly in individuals with predisposing factors. Healthcare providers need to be vigilant about this potential adverse effect and closely monitor renal function and serum bicarbonate levels in patients initiating or receiving high doses of zonisamide.

#### 4.3.4 New potential safety signals

This study identified several new potential safety signals associated with the drug ZNA. Among the top 30 risk signals based on adverse drug reaction (ADR) strength, ten were not previously listed in the drug’s specifications, including perinephritis, myoclonic dystonic epilepsy, ovarian granulosa vesicular cell tumor, positive serology for human herpesvirus 6, and positive lymphocyte stimulation test. Additionally, two of the top 30 risk signals based on ADR frequency were not documented in the drug’s instructions, namely infectious pneumonitis and over-the-counter use. These findings suggest that these adverse reactions may be linked to ZNA and should be further investigated in future studies and clinical treatments.

In our study, three cases of perinephritis were found and the signal was strong. The mechanism of perinephritis may be related to urinary tract obstruction caused by ureteral stones[20], and zonisamide increases the incidence of renal stones, which can lead to urinary tract obstruction causing hydronephrosis, and can also produce perirenal inflammation. In patients with hydronephrosis, increased pressure in the urinary tract leads to extravasation of fluid causing subperinephric fluid and oedema of the perinephric and pararenal interstitial structures. Although little is known, the associated risks should not be ignored. When patients present with symptoms of urinary tract infection such as low back pain, fever, haematuria, etc., CT examination should be selected to determine whether they have perinephritis.

This study also found 3 cases of ovarian granulosa cell tumor, all of which were adolescent females and were reported by physicians. There is no literature on the relationship between zonisamide and ovarian granulosa cell tumor. Combined with the epidemiology and risk factors of ovarian cell tumor[21], we analyzed that patients may have risk factors such as obesity or family history of ovarian cancer, and Zonisamide is not causally related.

Previous literature has reported that persistent latent infection and reactivation of HHV-6 could be linked to the development and progression of certain diseases, particularly autoimmune diseases like Drug-induced hypersensitivity syndrome [22]. Reactivation of HHV-6 is a common feature in DIHS and is associated with disease severity[23]. It has been proposed that ZNS might generate hydroxylamine, a metabolite that cannot be detoxified in DIHS-susceptible populations, leading to cytotoxic and immunological events that trigger hypersensitivity reactions[24]. Studies have shown that anti-herpesvirus treatment with ganciclovir in DIHS patients can help prevent or reduce HHV6 reactivation[25].

## 5. Limitations

The data for this study were obtained from the FAERS database, which is a spontaneous reporting system, and therefore there are some limitations and biases in this study. Firstly, the reported adverse drug reactions may be related to the underlying disease being treated; or caused by other medications taken concurrently, which do not necessarily constitute a direct causal relationship with the reported drug, and there is a possibility of overestimation of these signals[26]. In the present study, signal detection and initial screening were performed only for a single drug, zonisamide, and no drug combinations were considered for signal assessment, which may record AEs caused by other antiepileptic drugs as adverse events for zonisamide, leading to an increase or overestimation of AEs. The detected ADR signals only indicate a statistical correlation between the drug and the ADR signals, and are only suggestive of the potential for AE risk, and their causality still needs to be further studied and assessed in the context of the relevant literature and clinics. Subgroup analysis is an effective means to discover different positive signals and improve the sensitivity of data mining, which can effectively control confounding bias, a problematic factor in spontaneous presentation systems [27]. Second, the FAERS database exists with diverse sources of information (e.g., pharmacists, patients, doctors, etc.), and data standardisation problems often occur, such as: duplication of data, inconsistency in the names of drugs and adverse reactions, etc., and problems such as omission, misreporting, and missing information, which lead to reporting bias with the possibility of underestimation of signals.

## 6. Conclusion

This study utilized the ROR and BCNP combination to detect post-marketing adverse reactions of ZNS by analyzing the FAERS database. The systems with the highest frequency of PT signals were psychiatric and neurological disorders. While many of these adverse reactions were previously documented in prescribing information, some notable concerns were identified, including urinary stones, oculomucocutaneous syndromes, and DIHS. Additionally, potential new signals like HHV-6 were also observed. The prevalence of severe illnesses among ZNS adverse reactions underscores the importance of remaining vigilant and ensuring rational medication use in clinical settings.We emphasize that the results of this study, which represent purely statistical associations, do not imply a true cause-and-effect relationship; therefore, further research, including prospective cohort studies, is necessary for confirmation.

## Data Availability

All adverse event case files are available from the FAERS database (https://fis.fda.gov/extensions/FPD-QDE-FAERS/FPD-QDE-FAERS.html).

## Notes

**Funding** This study was supported by National Natural Science Foundation of China Youth Science Fund Project (Grant numbers: 81901324)

### Competing Interest Statement

The authors have declared no competing interest.

